# A novel molecular subtype of hepatocellular carcinoma based on the tumor purity and tumor microenvironment-related polygenic risk scores

**DOI:** 10.1101/2022.02.13.22270882

**Authors:** Yan Lin, Rong Liang, Xing Gao, Ziqin He, Lu Lu, Min Luo, Qian Li, Xiaobo Wang, Yongqiang Li, Guobin Wu, Xiaoling Luo, Jiazhou Ye

## Abstract

**Purpose:** The purpose of the present study was to use malignant cell-related and tumor microenvironment (TME)-related molecules to develop a novel molecular subtype of hepatocellular carcinoma (HCC).

**Methods:** The tumor purity (TP)-related and TME-related genes were identified and separately used to construct the TP-related and TME-related polygenic risk score (PRS). According to the two PRSs, we developed the TP-TME risk classification which was validated in two external data sets from The Cancer Genome Atlas Program and International Cancer Genome Consortium database. We also performed functional enrichment and drug repositioning analysis to reveal the potential biological heterogeneity among different subtypes.

**Results:** The three TP-TME risk subtypes of HCC had significantly different prognosis and biological characteristics. The TP-TME low risk subtype had the best prognosis and was characterized by well-differentiated, the TP-TME high risk subtype had the worst prognosis and was characterized by aberrant activation of TGFβ and WNT pathways, and the TP-TME high risk subtype had the moderate prognosis and was characterized by exhibited activated MYC targets and proliferation-related gene sets. These three TP-TME risk subtypes may respond differently to immunotherapy (e.g., immune checkpoint inhibitors and chimeric antigen receptor-modified T cells) or other drug therapies.

**Conclusion:** By combining the TP-related PRS and TME-related PRS, we proposed and validated the TP-TME risk subtyping system to divide patients with HCC into three subtypes with distinct biological characteristics and prognoses. These findings highlight the significant clinical implications of the TP-TME risk subtyping system and provide potential personalized immunotherapy strategies for HCC.

## INTRODUCTION

Hepatocellular carcinoma (HCC) is the most common primary liver cancer and ranks as the sixth most common neoplasm and the third leading cause of cancer death.(1) The development of HCC is a complex multistep process that involves the accumulation of somatic genomic alterations in driver genes in addition to epigenetic modifications, which lead to its huge molecular heterogeneity.(2) The current systems for HCC staging or subtyping are mainly according to radiologic, serologic, and pathologic-based tumor burden evaluations.(3) However, a previous study(4) indicated that HCCs at the same stage have diverse molecular characteristics. Thus, it is imperative to propose more precise subtyping system for predicting prognosis and treatment effects. Given the advances in sequencing technology, several molecular subtyping systems were developed according to multi-omics features of HCC, nevertheless, the differences of the previous studies in technological platforms, preparation, and processing of samples make it difficult to explore a common method for typing HCC.(5) Thus, none of these molecular classifications so far are recommended to predict disease progression or prognosis.(6)

In addition, heterogeneous tumor microenvironment (TME) in HCC is also a crucial part of tumor heterogeneity. It is now increasingly accepted that tumor cells are not working alone but interact closely with the TME.(7) The heterogeneous TME affects tumor response to various treatments.(8) Targeting the TME was proposed as a strategy for removing obstruction to anticancer immune responses and immunotherapy.(8) Intriguingly, few molecular classifications of HCC have so far taken into account both the related molecules of malignant cells and TME. Thus, in the present study, we tried to develop a novel molecular classification for HCC according to the expression patterns of malignant cell (tumor purity)- and TME-related genes. These unique risk factor patterns may provide a new frame to study cancer heterogeneity.

## MATERIALS AND METHODS

### Data Processing

We screened the HCCDB database (http://lifeome.net/database/hccdb/home.html)(9) to find the candidate data sets. The inclusion criteria were as follows: 1) the data set included both gene expression profiles and prognosis of patients with HCC, 2) the number of patients with a survival of more than 30 days should be more than 100, 3) and the gene expression profile of the dataset should contain more than 10,000 genes. According to the inclusion criteria, three data sets (GSE14520_GPL3921, TCGA-LIHC, and LIRI-JP) were selected and downloaded from HCCDB database for our analyses. Data set GSE14520_GPL3921(10) included the gene expression profiles based on the GPL3921 platform containing 225 HCC and 220 tumor-adjacent liver tissue samples were utilized to develop our subtyping systems. TCGA-LIHC data set containing RNA sequencing (RNA-seq) data and clinical information of 356 HCC belongs to The Cancer Genome Atlas Program (https://www.cancer.gov/tcga), and LIRI-JP data set containing RNA-seq data and clinical information of 212 HCC from JP Project from International Cancer Genome Consortium (https://dcc.icgc.org/) were used to validated the subtyping systems. The workflow of the present was shown in **Figure 1**.

**Figure 1.**
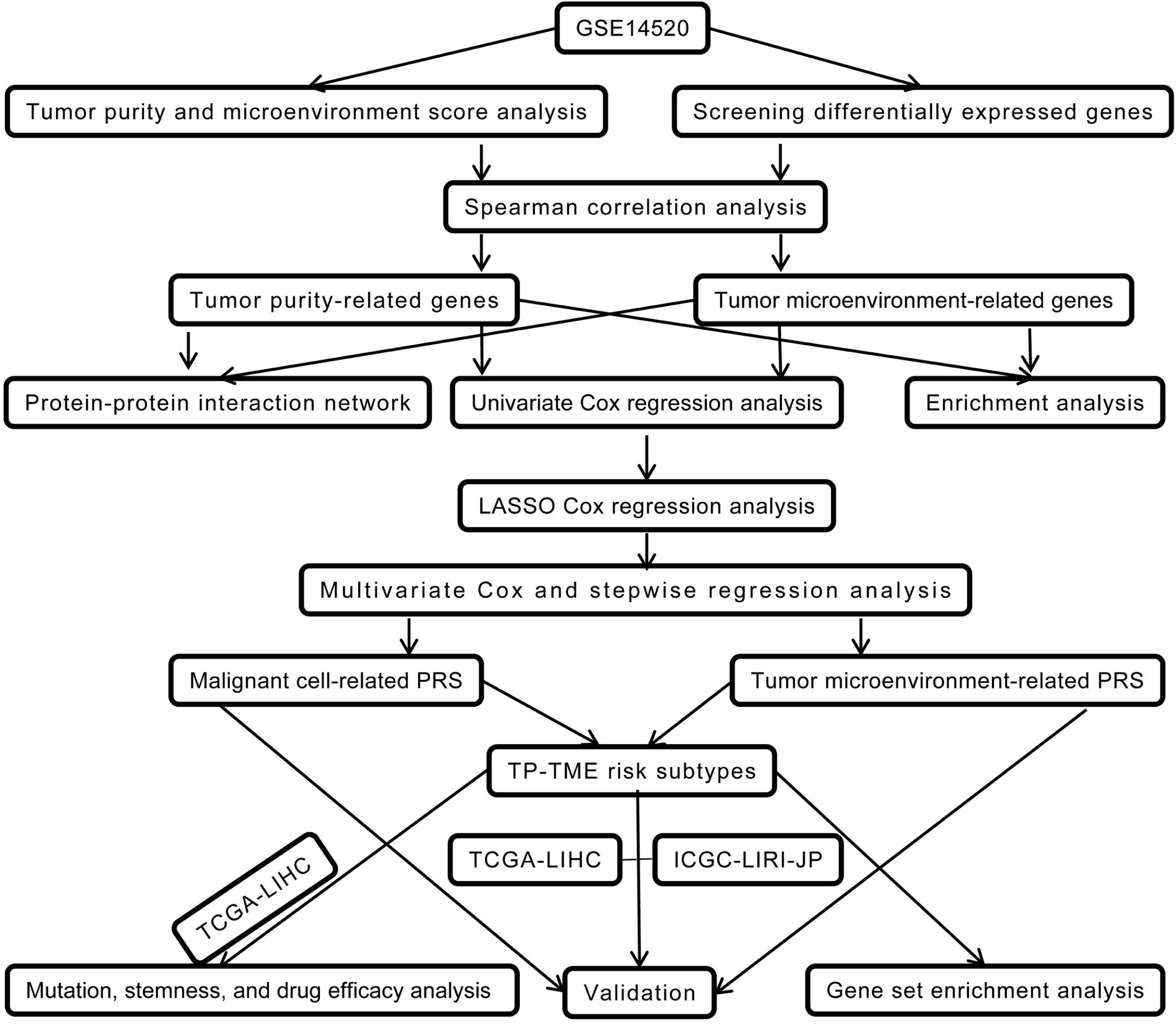
The workflow of the present study. **Abbreviation**: LASSO, least absolute shrinkage and selection operator; TP, tumor purity; TME, tumor microenvironment; TCGA-LIHC, The Cancer Genome Atlas-liver hepatocellular carcinoma; ICGC-LIRI-JP: Liver Cancer-RIKEN, JP.

### Calculation of tumor purity and TME score and identification of differentially expressed genes (DEGs) in GSE14520_GPL3921

The gene expression profiles of GSE14520_GPL3921 were firstly utilized to calculate tumor purity (TP) using the ESTIMATE package.(11) Then, GSE14520_GPL3921 was also used to calculate the TME score using the xCell tools (https://xcell.ucsf.edu/)(12) with the xCell gene signature. The DEGs in HCC compared to tumor-adjacent liver tissue were identified using limma package.(13) Genes with fold changes > 1.5 and P (adjusted by false discovery rate) value < 0.05 were considered significant.

### Normality test and correlation analysis

The tumor purity and TME score were separately performed Shapiro-Wilk test. Spearman or Pearson correlation analyses were performed to calculate the correlation between DEGs and TP and TME score. A DEG showed a positive correlation with TP and a negative correlation with TME score was considered a TP-related gene, while a DEG showed a negative correlation with TP and a positive correlation with TME score was considered a TME-related gene. In addition, the TME-related genes do not include mark genes of TME cells in xCell signature.

### Protein-protein interaction (PPI) networks

The PPI networks of TP-related and TME-related genes were obtained from STRING database (version 11.5)(14) to preliminarily reveal the crosstalks between the tumor cells and TME. The interactions with the high confidence (>0.7) were included in our present study and visualized using the Cytoscape software (version 3.8.0).(15)

### Development of the TP- and TME-related gene-based polygenic risk scores

Firstly, for developing the TP-related polygenic risk score (PRS), the overall survival (OS)-associated TP-related genes were identified using univariate Cox regression analysis. Secondly, the expression profiles of the OS-associated TP-related genes were used to carry out the least absolute shrinkage and selection operator (LASSO) Cox regression model analysis with leave-one-out cross validation using glmnet package.(16) The genes with non-zero coefficient were considered the optimal features and subjected to multivariate Cox regression and stepwise regression analysis. Then the TP-related PRS was developed as the formula: TP-related PRS = Σ (Expression_i_ * Coeffient_i_). Where the “Coeffient” and “Expression” represent the risk coefficient and expression of each gene in the multivariate cox regression and stepwise regression analysis, respectively. The TME-related PRS was also developed according to the same method as above.

### The TP-TME subtyping of HCC

The optimal cutoffs of the TP-related and TME-related PRS were identified using the surv_cutpoint function from survminer package (https://CRAN.R-project.org/package=survminer) to separately divide patients into high and low TP- and TME-related PRS groups. Each individual got a TP- and a TME-related PRS level, and we developed the TP-TME subtype according to the TP- and TME-related PRS levels. Patients possessing high TP- and TME-related PRS were considered as the high-risk subtype, those possessing low TP- and TME-related PRS were considered the low-risk subtype, and the remaining patients possessing a high TP-related and low TME-related PRS or a low TP-related and high TME-related PRS were considered the intermediate-risk subtype.

### Gene set enrichment analysis (GSEA)

In order to preliminarily reveal the biological characteristics of these three risk subtypes, we performed GSEA(17, 18) in the GSE14520_GPL3921 data set using the GSEA java software (http://www.gsea-msigdb.org/gsea/index.jsp). Hallmark gene sets and canonical pathway gene sets derived from the Kyoto Encyclopedia of Genes and Genomes (KEGG) pathway database were download from The Molecular Signatures Database (MSigDB)(18-20) and used as the reference gene sets. The threshold was set to nominal P (NOM P) value < 0.05 or FDR q value < 0.25.

### Analyses of gene mutation and stemness score

Gene mutation data of TCGA-LIHC data set were extracted from mutation annotation format (MAF) files using the *GDCquery_Maf* function in the “TCGAbiolinks” package.(21) Gene mutation frequencies of each risk subtype were visualized as a waterfall plot using the *oncoplot* function in the “TCGAbiolinks” package. The tumor mutational burden (TMB) of each sample was obtained from a previous study.(22) The stemness score(23) was calculated for each individual in the TCGA-LIHC data set using *TCGAanalyze_Stemness* function in the “TCGAbiolinks” package.

### Prediction of efficacy of therapy

Immunotherapy has achieved tremendous successes in treatment of various cancers,(24) including HCC.(25) Treatment of immune checkpoint inhibitors (ICIs) and chimeric antigen receptor-modified T cell (CAR-T) were currently the two most widely studied immunotherapies. The expression of immune checkpoints was associated with the efficacy of ICIs.(26) The therapeutic effect of CAR-T cells is related to the expression of target genes in the tumor cells. For the TP-TME risk subtypes, we compared the expression levels of two immune checkpoints (PDL1 and CTLA4) and five antigens (CD133, EPCAM, GCP3, MSLN, and MUC1)(27) to predict the potential response to these treatments. In addition, we performed drug repositioning analysis for the high-risk subtype using the PATHOME-Drug (http://statgen.snu.ac.kr/software/pathome/) web tools.

### Statistical analysis

In our present study, unless otherwise stated, all these analyses were performed in R (version 4.0.2). We identified DEGs using unpaired t-tests provided by limma package. Shapiro-Wilk test was used for the nrmality test. Time-dependent receiver operating characteristic curve (tROC) analysis was carried out using the tROC package.(28) Kaplan–Meier survival curves for overall survival (OS) and progression free survival (PFS) were compared in different subtypes using the log-rank method in the “survival” package (https://CRAN.R-project.org/package=survival) and “survminer” package (https://CRAN.R-project.org/package=survminer). Intergroup differences in continuous variables were assessed for significance using Wilcoxon, Kruskal–Wallis, or unpaired t-tests. All tests were two-sided and unless otherwise stated, we set P value < 0.05 to be statistically significant.

## RESULTS

### The biological functions and interactions of tumor purity (TP)-related and TME-related genes

We identified a total of 2263 DEGs in HCC compared to tumor-adjacent liver tissue (**Figure 2A**). Both the TP and TME score showed non-normal distribution with Shapiro-Wilk P < 0.001. A total of 451 TP-related and 121 TME-related genes were identified by Spearman correlation analysis, and the bidirectional hierarchical clustering showed the expression patterns of them could basically distinguish HCC and tumor-adjacent liver tissue (**Figure 2B**). Unsurprisingly, the TP-related genes are mainly involved in cancer-related gene ontology terms or pathways **(Figure 2C**), such as cell cycle and mismatch repair; while the TME-related genes are mainly involved in immune system (**Figure 2D**). The PPI networks of the TP-related and TME-related genes contain 342 nodes and 1177 edges (**Figure 2E**). Red indicates upregulated and blue indicates downregulated in the metastasis group, while circular nodes represent the TP-related genes and rhombic nodes represent the TME-related genes.

**Figure 2.**
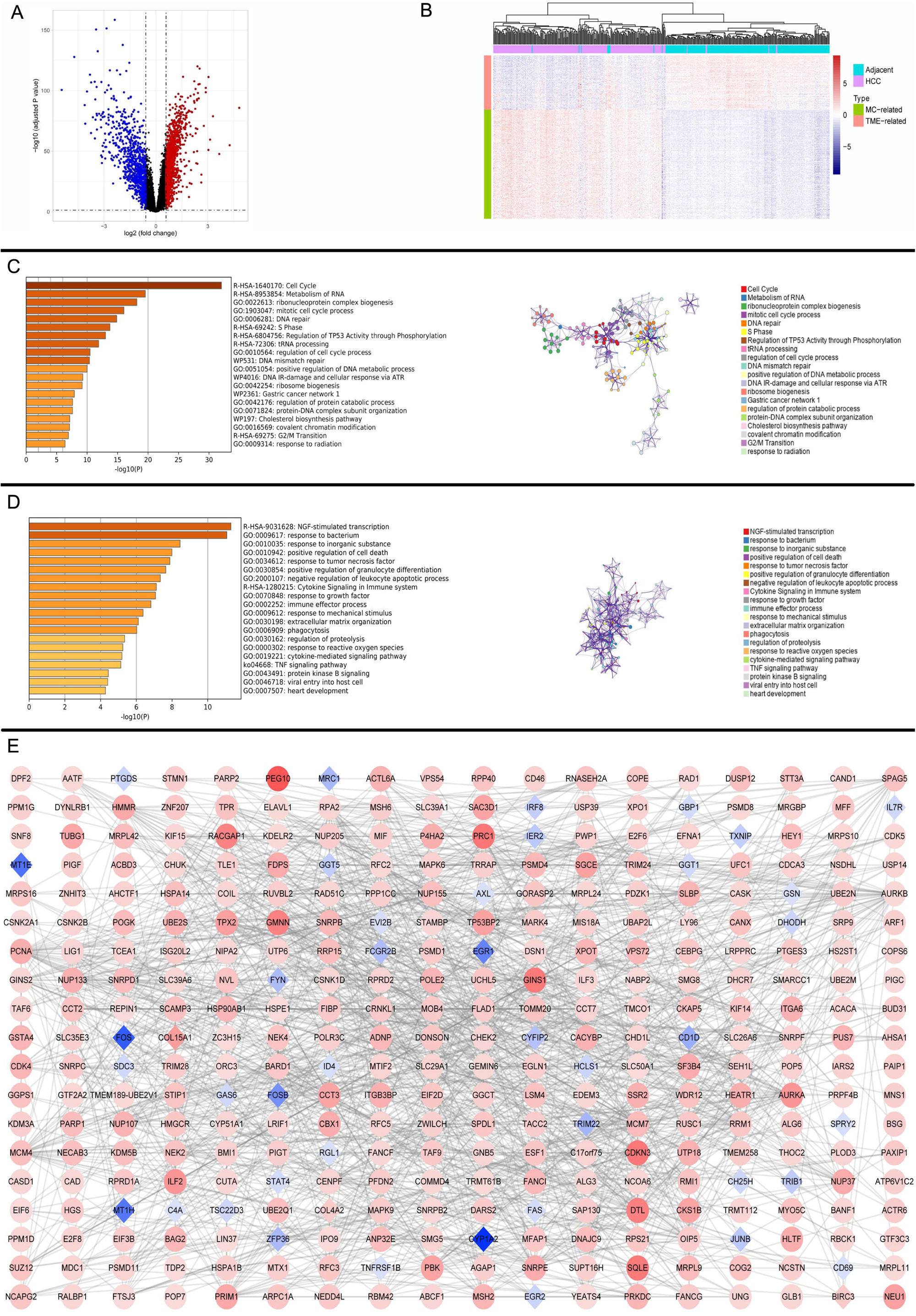
The identification, enrichment analysis, and protein-protein interaction networks of the tumor purity-related genes and tumor microenvironment-related genes. **(A)** The volcano plot of the differentially expressed gene analysis; **(B)** Hierarchical clustering showed the expression patterns of tumor purity (TP)-related and tumor microenvironment (TME)-related genes basically distinguish hepatocellular carcinoma (HCC) and tumor-adjacent liver tissues. **(C)** Functional enrichment analysis for the TP-related genes. *Left panel:* GO terms and pathways involving TP-related genes, and *right panel*: interactions among the GO terms and pathways. **(D)** Functional enrichment analysis for the TME-related genes. *Left panel:* GO terms and pathways involving TME-related genes, and *right panel*: interactions among the GO terms and pathways. **(E)** protein-protein interaction networks of the TP-related genes and TME-related genes. The red represents up-regulated, and the blue represents down-regulated in hepatocellular carcinoma. The circle nodes were TP-related genes, and the diamond nodes were TME-related genes. **Abbreviation:** TME, tumor microenvironment; TP, tumor purity; HCC, hepatocellular carcinoma; GO, gene ontology.

### The TP-TME risk subtyping is a robust prognosis prediction system

Fifty TP-related genes were identified as OS-associated genes, twenty-two of them possess non-zero coefficient (**Figure 3A**), and eleven genes (*ALG6, ATP5MF, CNIH4, ESM1, HEY1, LANCL1, P2RX4, PEX11B, POP7, RCN2*, and *XPO1*) were used to generate the TP-related polygenic risk score (PRS) (**Supplementary Table 1**). The TP-related PRS was significantly associated with overall survival (OS) with P < 0.0001, Hazard Ratio (HR) = 2.718 (95%CI for HR = 2.147-3.442) and the area under curve (AUC) of tROC analysis was stably around 0.8 (**Figure 3B**). The HCC patients with high TP-related PRS showed shorter OS than those with low TP-related PRS (P < 0.0001) (**Figure 3C**). In the TME-related genes, twelve genes were identified as OS-associated genes, ten of them possess non-zero coefficient (**Figure 3D**), and seven genes (*ALDH1B1, CTSC, GUCY1A1, MRC1, SPRY2, TARP*, and *TRIM22*) were used to generate the TME-related PRS (**Supplementary Table 2**). The TME-related PRS was also significantly associated with OS with P < 0.0001, Hazard Ratio (HR) = 2.718 (95%CI for HR = 1.978-3.735) and the AUC of tROC analysis was 0.7-0.8 (**Figure 3E**). The HCC patients with high TME-related PRS showed shorter OS than those with low TP-related PRS (P < 0.0001) (**Figure 3F**). Our TP-TME risk subtype was generated based the two PRSs, and 34, 52, and 123 patients with HCC were divided into high-, intermediate-, and low-risk subtypes, respectively. Cases defined as high-risk subtype had the best poor survival while those in low-risk subtype had the best survival, and the intermediate-risk cases had a better prognosis than high-risk subtype and worse than low-risk subtype (**Figure 3G**). A similar trend was also observed in progression free survival (PFS) (**Figure 3H**). Furthermore, the TP-TME risk subtyping system showed independent to some routine clinicopathological features (**Figure 3I**). As it was in the GSE14520_GPL3921 data set, the TP-related PRS and the TME-related PRS in the TCGA-LIHC and LIRI-JP data sets were calculated according to the abovementioned formula, respectively. We found similar results in the two validation sets. Briefly, the TP-related PRS (**Figure 4A** for TCGA-LIHC and **Figure 5A** for LIRI-JP) and the TME-related PRS (**Figure 4B** for TCGA-LIHC and Figure 5B for LIRI-JP) were significantly associated with OS, the TP-TME risk subtyping system was associated with OS (**Figure 4C** for TCGA-LIHC and **Figure 5C f**or LIRI-JP) and PFS (**Figure 4D** for TCGA-LIHC). The prognostic value of the TP-TME risk subtyping system was independent to the routine clinicopathological features (**Figure 4E** for TCGA-LIHC and **Figure 5D** for LIRI-JP). Collectively, the TP-TME risk subtyping is a robust prognosis prediction system.

**Figure 3.**
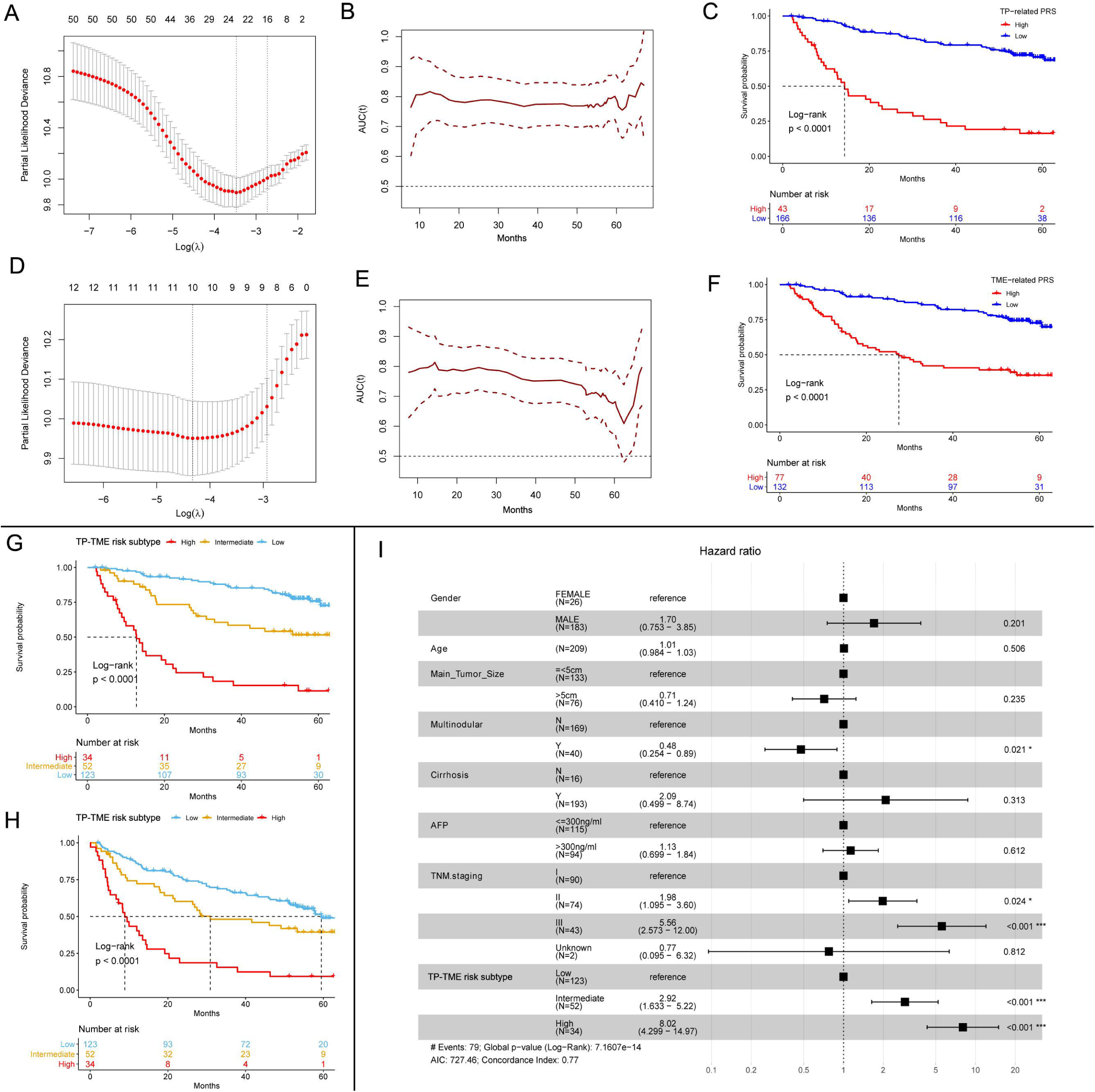
The development of TP-TME risk subtypes in GSE14520_GPL3921 data set. **(A)** twenty-two TP-related genes had non-zero coefficients in the LASSO Cox regression model analysis; **(B)** Time-dependent ROC curve analysis for the TP-related PRS; **(C)** HCC with high TP-related PRS had shorter overall survival than those with low TP-related PRS. **(D)** Ten TME-related genes had non-zero coefficients in the LASSO Cox regression model analysis; **(E)** Time-dependent ROC curve analysis for the TME-related polygenic risk score; **(F)** HCC with high TME-related PRS had shorter overall survival than those with low TME-related PRS. **(G)** There are significantly different overall survivals between the three subtypes in the TP-TME risk subtypes. **(H)** There are significantly different progression free survivals between the three subtypes in the TP-TME risk subtypes. **(I)** The TP-TME risk subtype system was proved to be an independent prognostic factor, after adjusting for other clinicopathological characteristics. **Abbreviation:** LASSO, least absolute shrinkage and selection operator; TP, tumor purity; TME, tumor microenvironment; HCC, hepatocellular carcinoma; ROC, receiver operating characteristic; PRS, polygenic risk score.

**Figure 4.**
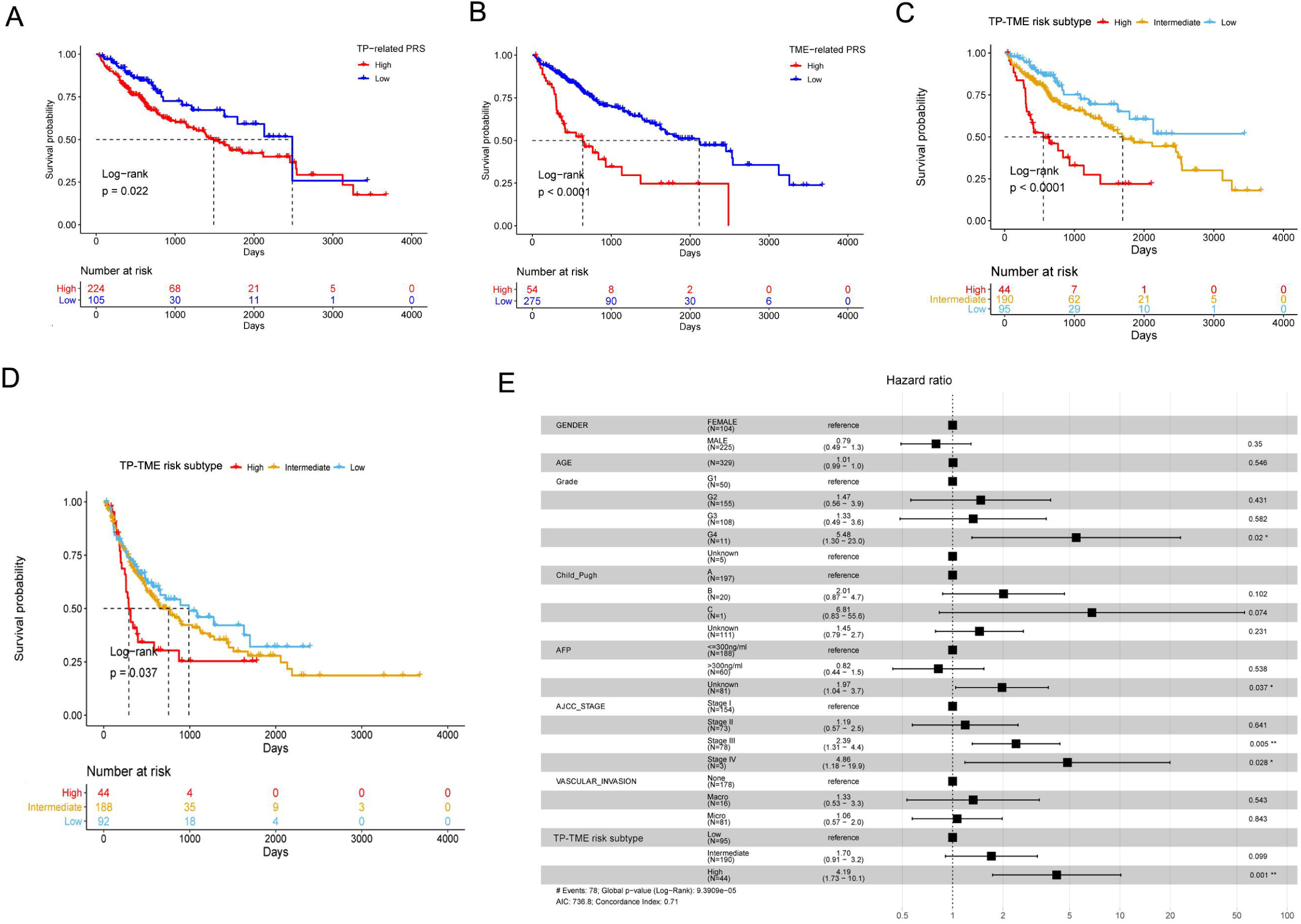
The validation of TP-TME risk subtypes in TCGA-LIHC data set. **(A)** HCC with high TP-related PRS had shorter overall survival than those with low TP-related PRS. **(B)** HCC with high TME-related PRS had shorter overall survival than those with low TME-related PRS. **(C)** There are significantly different overall survivals between the three subtypes in the TP-TME risk subtypes. **(D)** There are significantly different progression free survivals between the three subtypes in the TP-TME risk subtypes. **(E)** The TP-TME risk subtype system was proved to be an independent prognostic factor, after adjusting for other clinicopathological characteristics. **Abbreviation:** TP, tumor purity; TME, tumor microenvironment; PRS, polygenic risk score; AFP, alpha fetoprotein.

**Figure 5.**
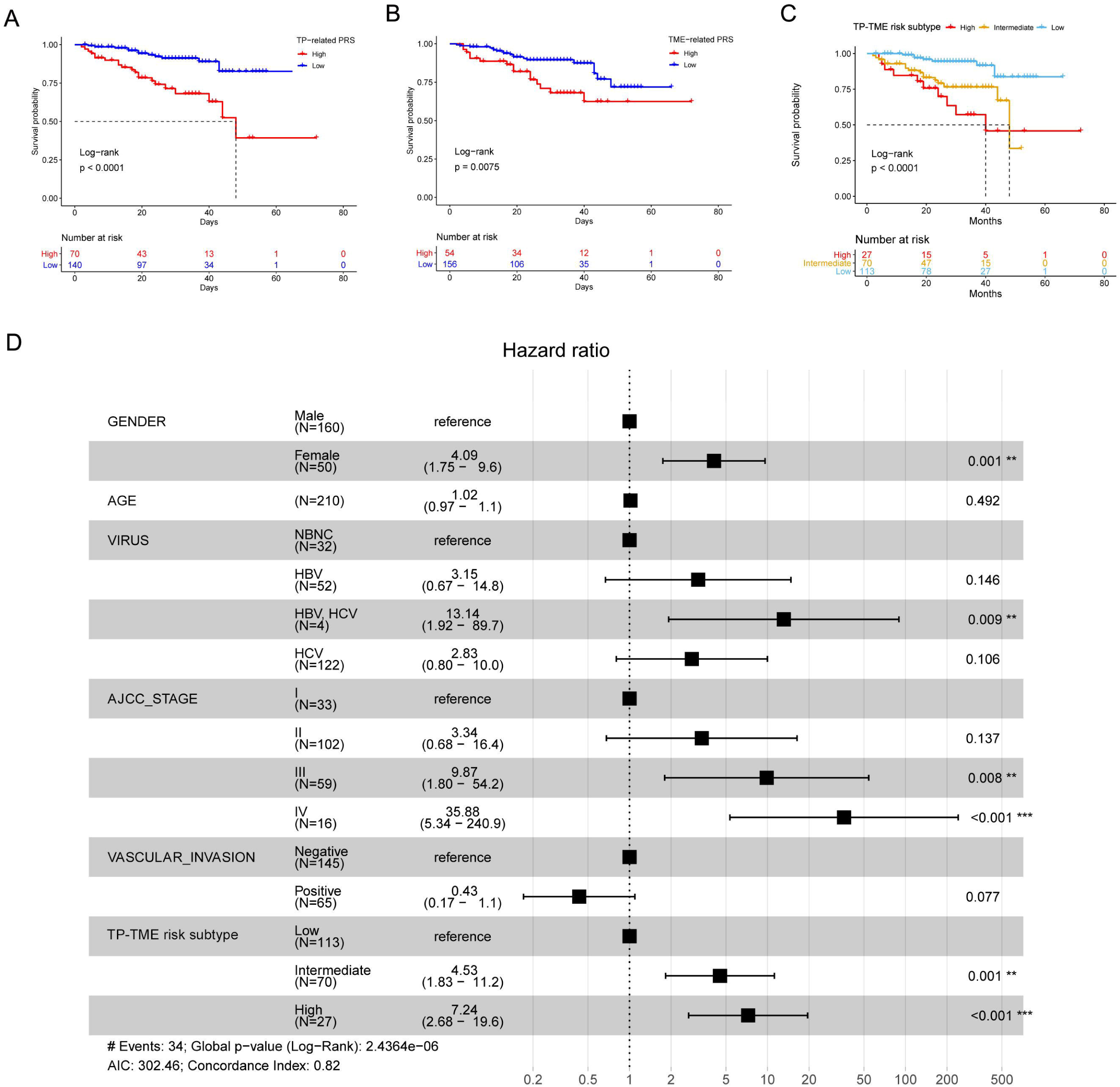
The validation of TP-TME risk subtypes in LIRI-JP data set. **(A)** HCC with high TP-related PRS had shorter overall survival than those with low TP-related PRS. **(B)** HCC with high TME-related PRS had shorter overall survival than those with low TME-related PRS. **(C)** There are significantly different overall survivals between the three subtypes in the TP-TME risk subtypes. **(D)** The TP-TME risk subtype system was proved to be an independent prognostic factor, after adjusting for other clinicopathological characteristics. **Abbreviation:** TP, tumor purity; TME, tumor microenvironment; PRS, polygenic risk score; NBNC, no hepatitis B virus and no hepatitis C virus; HBV, hepatitis B virus; HCV, hepatitis C virus; AJCC, American Joint Committee on Cancer.

### The subtype-specific curated gene sets of the TP-TME risk subtypes

Compared to the TP-TME intermediate- and high-risk subtypes, the liver function-related hallmark (**Figure 6A**) and metabolism-related Kyoto Encyclopedia of Genes and Genomes (KEGG) (**Figure 6D**) gene sets were significantly enriched in the TP-TME low-risk subtype. This suggests the TP-TME low-risk subtype HCC were well-differentiated. The TP-TME intermediate-risk subtype was charactered by enriching transcription factor *E2F* and *MYC* targets (**Figure 6B**) and cell cycle pathways (**Figure 6E**), while the TP-TME high-risk subtype was charactered by enriching hypoxia and Wntβ catenin signaling (**Figure 6C**), and Notch signaling pathway and TGFβ signaling pathway (**Figure 6F**). These results indicated that there is significant biological heterogeneity between these three subtypes.

**Figure 6.**
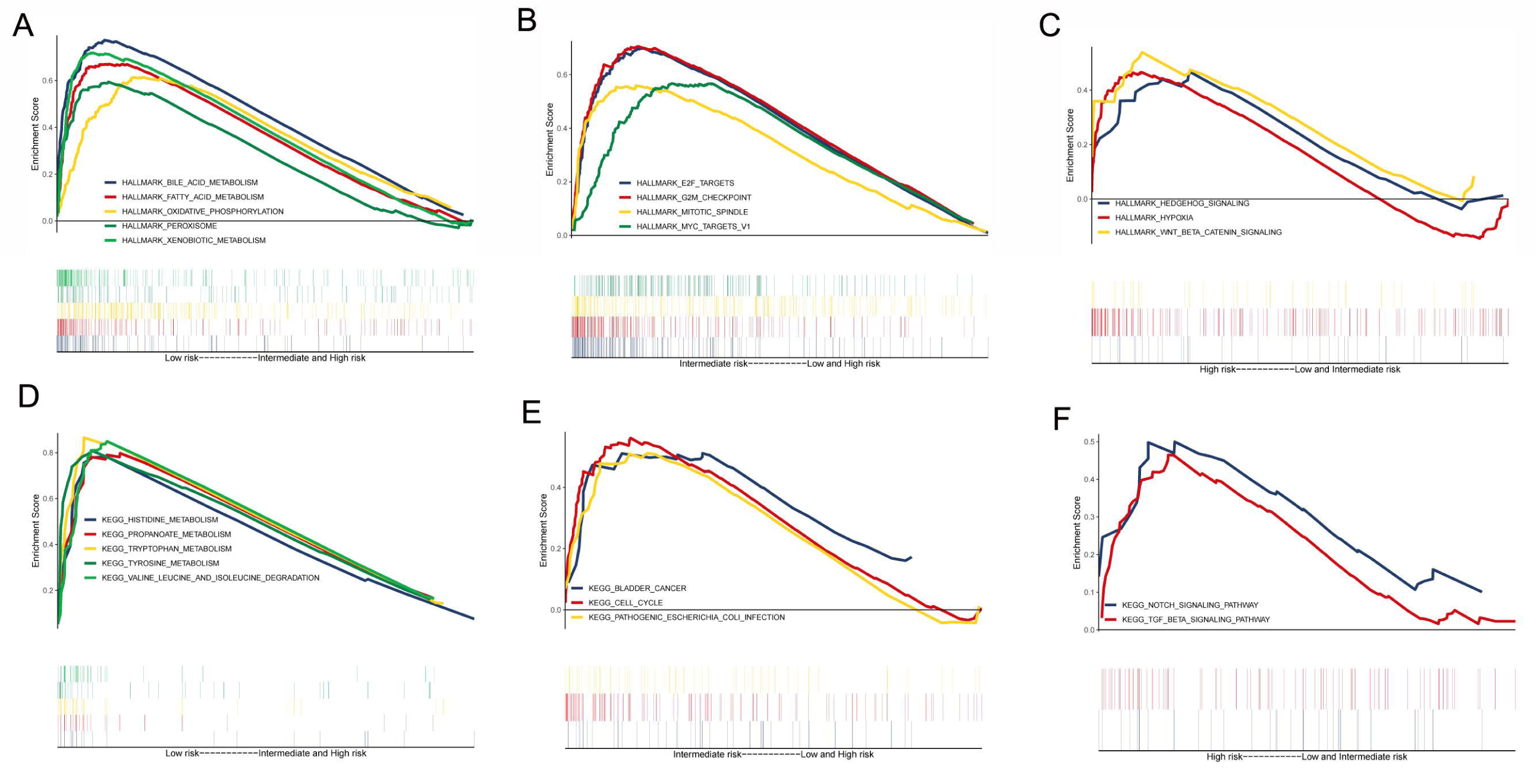
Gene set enrichment analysis for identifying the subtype-specific curated gene sets of the TP-TME risk subtypes. The hallmark gene sets enriched in the **(A)** TP-TME low-risk subtype, **(B)** TP-TME intermediate-risk subtype, and **(C)** TP-TME high-risk subtype. The Kyoto Encyclopedia of Genes and Genomes (KEGG) pathway gene sets enriched in the **(D)** TP-TME low-risk subtype, **(E)** TP-TME intermediate-risk subtype, and (F) TP-TME high-risk subtype.

### These three TP-TME risk subtypes may respond differently to immunotherapy

The **Figures 7A-C** show the top 30 mutation genes in the low-, intermediate-, and high-risk subtypes, respectively. However, few known drugs so far targeted these genes for HCC. The tumor mutational burden (TMB) in HCC was low, and no significantly different among these three subtypes (**Figure 7D**). This suggests TMB may not be an efficient biomarker for selecting patients with HCC for ICI treatment. In addition, our analysis also found no significant difference in the stemness score among these three risk subtypes (**Figure 7E**). Though the CD274 (also known as PDL1) expression of the three risk subtypes show no significantly differentially (**Figure 7F**), the expressions of CTLA4 (**Figure 7G**) and PDCD1 (also known as PD1) (**Figure 7H**) were higher in the intermediate-risk subtypes than the low-risk subtypes. Thus, the intermediate-risk subtype may possess a higher response rate to treatment with ICIs more than the low-risk subtype. In addition, the three TP-TME risk subtypes have distinct expression levels of the five cancer antigens which were used as targets in the chimeric antigen receptor-modified T cell (CAR-T) therapy. The GPC3 express higher in the intermediate-risk subtype than the low-risk subtype (**Figure 7I**), the MSLN express higher in the intermediate- and high-risk subtypes than the low-risk subtypes (**Figure 7J**). Impressively, the high-risk subtypes have highest expression of MUC1 (**Figure 7K**), EPCAM (**Figure 7L**), and PROM1 (**Figure 7M**). Thus, theoretically, the three TP-TME risk subtypes may respond differently to corresponding CAR-T therapies.

**Figure 7.**
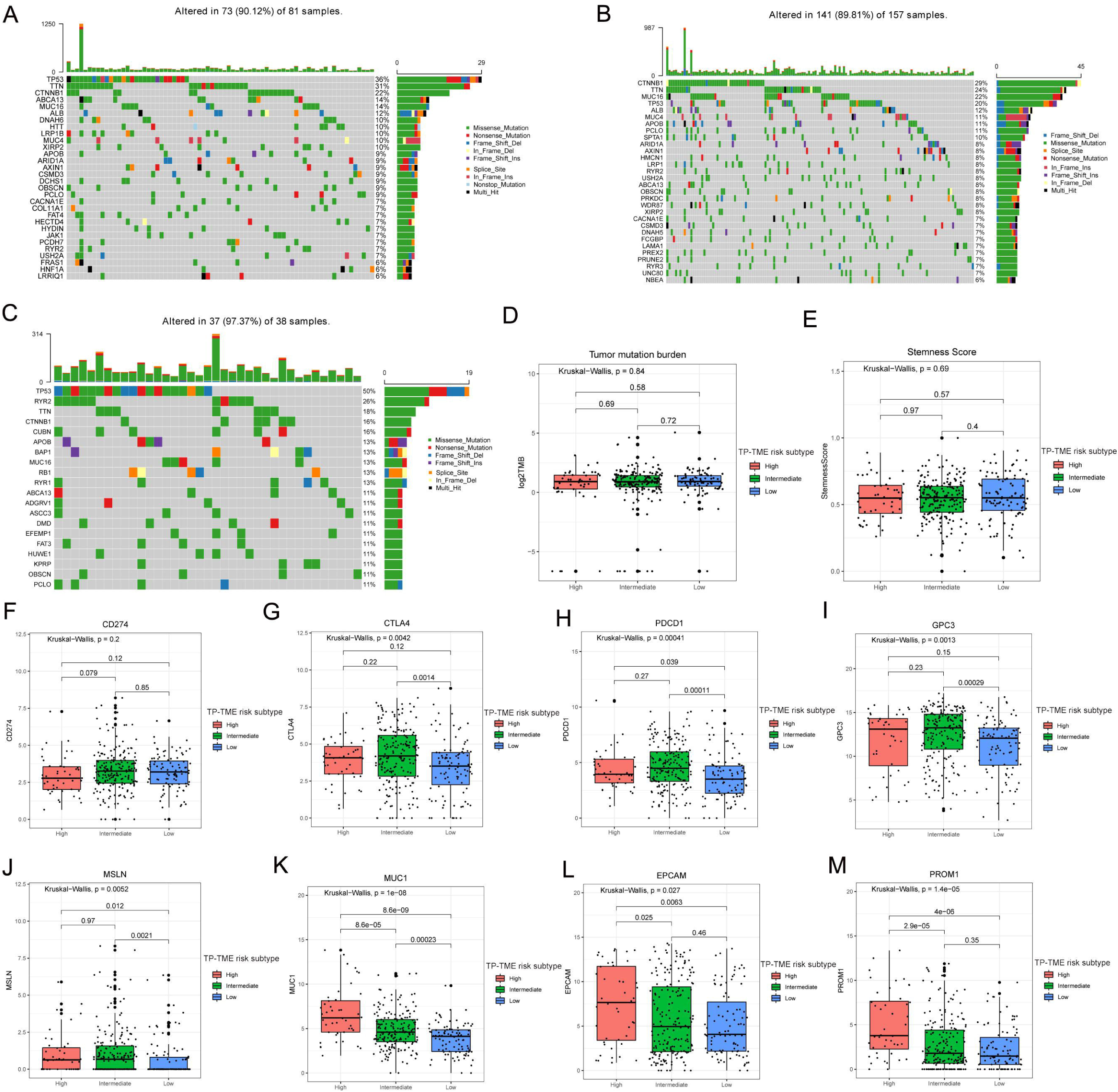
Mutation, stemness, and immunotherapeutic efficacy analysis. **Top 30 mutant genes in the (A)** TP-TME low-risk subtype, **(B)** TP-TME intermediate-risk subtype, **(C)** and TP-TME high-risk subtype. **(D)** Tumor mutation burden in the three TP-TME high-risk subtypes. **(E)** Stemness score in the three TP-TME high-risk subtypes. The expression of (F) CD274, (G) CTLA4, **(H)** PDCD1, (I) GPC3, **(J)** MSLN, (K) MUC1, **(L)** EPCAM, and **(M)** PROM1 in the three TP-TME high-risk subtypes.

### Potentially effective drugs for the high-risk subtypes

Many agents which tried to treat HCC evaluated in phase 3 trials got the disappointing results,(1) the response is usually observed in a small subset of individuals. Through the PATHOME-Drug analysis, we constructed the drug-target networks to identify the potentially effective drugs for the high-risk subtypes (**Figure 8**). The potentially effective drugs include recommended agents such as sorafenib, regorafenib, and cabozantinib, and some drugs which are used to treat other diseases (**Supplementary Table 3**).

**Figure 8.**
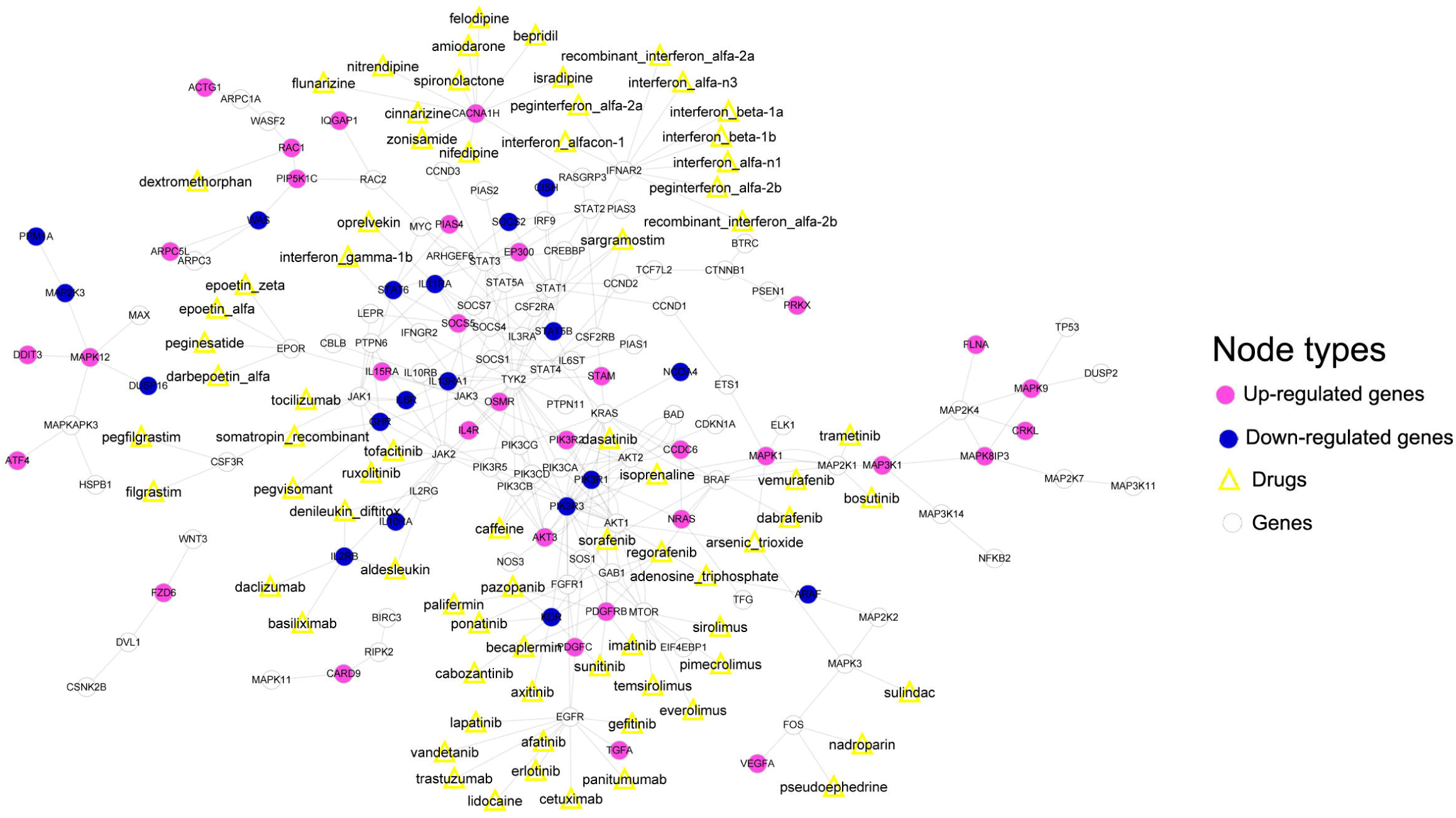
Drug-target networks for potentially effective drugs for the TP-TME high-risk subtypes. The red represents up-regulated, and the blue represents down-regulated in the TP-TME high-risk subtypes.

## DISCUSSION

The heterogeneity of HCC is attributed to the presence of various etiologies, such as infections of virus or parasitic, chemical carcinogens, cigarette smoking, excess alcohol intake, and dietary factors.(1) Accordingly, the host’s response to various pathogenic factors leads to the formation of their own unique microenvironment. One hypothesis is that different TMEs and pathogenic factors may evoke distinct molecular alteration to independently initiate HCC progress, which results in extensive inter-tumor molecular heterogeneity. One of the essential efforts for improving the poor outcome of HCC is to provide a subtyping system that is capable of accurately defining tumor risk subtypes, each displaying unique molecular characteristics linked to potentially druggable driver genes in order to provide personalized treatment choices based on the subtyping system. Though many efforts, mainly focusing on the malignant cells, were paid to elaborate the inter-tumor heterogeneity and propose various single- or multi-omics-based molecular typing systems,(5, 29) their effectiveness remain limited for providing precision treatment. In this scope, given that the crucial role of TME in cancers is confirmed,(30) TME-related molecules should be contributed to the subtyping for HCC. Another challenge of previous molecular typing methods is cost effectiveness, due to they need hundreds of genes or even multiple omics data types.

In our present study, we firstly identified the related genes of TP and TME and subsequently generated a TP-related PRS and a TME-related PRS according to the expression patterns of these types of genes, and furtherly proposed a novel risk subtyping that could successfully divide patients with HCC into three risk subtypes. Similar to other molecular typing systems,(31-34) our subtypes have distinct prognosis and were validated in two independent external data sets. As reviewed by Wu et al.(5) a few subtypes were repeatedly uncovered in different studies, indicating that different HCC subtypes derived from different omics technologies may share common molecular characteristics. Our TP-TME low-risk subtype may be well-differentiated and enrich gene sets related to liver function (e.g., bile acid metabolism), similar to the Hoshida’s S3 subtype(35) and TCGA’s ICluster2. The TP-TME intermediated-risk subtype exhibited activated MYC targets and proliferation-related gene sets (e.g., cell cycle and G2M checkpoint), corresponding to the Hoshida’s S2 subtype and Chaisaingmongkol et al.’s C1 subtype.(36) The TP-TME high-risk subtype characterized by aberrant activation of TGFβ and WNT pathways, displayed multiple similarities with Hoshida’s S1 subtype and Kurebayashi et al.’s immune-high subtype.(37) Compared with these typing methods, low cost is the potential advantage of our TP-TME risk subtyping system. In addition, though the further study was required, we also proposed the potential immunotherapy and drugs for the high-risk subtype, which may help for decision-making in clinical practice.

Unsurprisingly, some of these the candidate eleven TP-related and seven TME-related genes have been found to be associated with HCC or other types of cancers in previous studies. *ESM1* was found as a biomarker of macrotrabecular-massive HCC.(38) *HEY1* plays a critical role in hypoxia-related regulation of mitochondrial activity in HCC.(39) *RCN2* can enhance HCC proliferation via modulating the EGFR-ERK pathway.(40) The interactions between *CTSC* and TNF-α/p38 MAPK signaling pathway are associated with proliferation and metastasis in HCC.(41) LANCL1 was reported that can protect prostate cancer cells from oxidative stress.(42) XPO1-dependent nuclear export was proposed as a target for cancer therapy.(43) However, the associations of some TP-related or TME-related genes and HCC have been not noted in the previous study. According to our current analysis, we provide potential candidate molecules for further research on HCC.

Although our current study provided a novel molecular classification system, it has some noted limitations. Firstly, the roles of some candidate genes in HCC remain elusive, it is not clear whether these genes are causal or merely prognostic markers for HCC. Secondly, the TP-TME risk subtyping system was generated from a retrospective analysis, it should be validated or improved by the prospective trials before being used in clinical practice.

## CONCLUSIONS

In conclusion, by combining separately constructed TP-related PRS and TME-related PRS, we proposed and validated a novel molecular classification system, the TP-TME risk subtyping system, to divide patients into three subtypes with distinct biological characteristics and prognosis. These findings highlight the significant clinical implications of the TP-TME risk subtyping system and provide potential personalized immunotherapy strategies for patients with HCC.

## Data Availability

The datasets used during the current study are available in the HCCDB repository,

http://lifeome.net/database/hccdb/home.html

## ACKNOWLEDGMENTS

Not available

## AUTHORS CONTRIBUTIONS

YJZ, LXL, and WGB designed the study and revised the manuscript. LY and LR performed the analyses and wrote the manuscript. GX, HZQ, LL, LM, LQ, WXB, and LYQ assisted with analyzing the data and writing the manuscript. All authors read and approved the final manuscript.

## DATA AVAILABILITY

The datasets used during the current study are available in the HCCDB repository, [http://lifeome.net/database/hccdb/home.html]

## CODE AVAILABILITY

The code used during the current study is available from the corresponding author upon request.

## ETHICS APPROVAL

Not available.

## CONSENT TO PARTICIPATE

Informed consent was obtained from all individual participants included in the study.

## CONSENT TO PUBLISH

Not available.

## Figure legends

**Supplementary Table 1 The overall survival-associated tumor purity-related genes in hepatocellular carcinoma**

**Supplementary Table 2 The overall survival-associated tumor microenvironment-related genes in hepatocellular carcinoma**

**Supplementary Table 3 Potentially effective drugs and relevant targets for the high-risk subtypes**

